# Methodological approach to sleep state misperception in insomnia disorder: comparison between multiple nights of actigraphy recordings and a single night of polysomnography recording

**DOI:** 10.1101/2023.07.25.23293105

**Authors:** Antonia Maltezos, Aurore A. Perrault, Nyissa A. Walsh, Emma-Maria Phillips, Kirsten Gong, Lukia Tarelli, Dylan Smith, Nathan E. Cross, Florence B. Pomares, Jean-Philippe Gouin, Thien Thanh Dang-Vu

## Abstract

**Rationale:** Sleep state misperception (SSM) represents the discrepancy between objectively recorded and subjectively perceived measures of sleep, including sleep onset latency (SOL), sleep duration (TST) and wake duration (WASO). The severity of SSM is higher in insomnia disorder (INS) compared to other populations. SSM is typically assessed in-lab during one night with polysomnographic (PSG) recording or at-home with actigraphy recordings over multiple days, both complemented by subjective sleep reports. Both methods of data collection have their specific strengths and weaknesses, and provide sleep measures that may differ, especially in individuals with sleep disorders. The extent to which the methods and environment of data collection impact measures of sleep misperception remains unclear. This study aimed at providing a comprehensive assessment of SSM in INS and good sleepers (GS) by comparing recordings performed for one night in-lab (PSG and night review) and during several nights at-home (actigraphy and sleep diaries).

**Methods:** Fifty-seven INS and 29 GS wore an actigraphy device and filled a sleep diary for two weeks at-home. They subsequently completed a PSG recording and filled a night review the next morning in-lab. Sleep perception index (subjective/objective × 100; in %) of SOL, WASO and TST were computed and compared between methods and groups.

**Results:** We found that GS and INS exhibit opposite patterns of sleep misperception. GS displayed a tendency to overestimate TST and WASO but correctly perceived SOL. The degree of misperception was similar across methods within the GS group. In contrast, INS underestimated their TST and overestimated their SOL both in-lab and at-home, yet the severity of misperception (i.e., degree of mismatch) of SOL was larger at-home than in-lab. Finally, INS overestimated WASO only in-lab while correctly perceiving it at-home. While only the degree of TST misperception was stable across methods in INS, misperception of SOL and WASO were dependent on the method used.

**Conclusions:** We found that GS and INS exhibit opposite patterns and severity of sleep misperception. While the degree of misperception in GS was similar across methods, we found that only sleep duration misperception was reliably detected by both in-lab and at-home methods in INS. Our results highlight that, when assessing sleep misperception in individuals with insomnia disorder, the method of data collection should be carefully considered in relation to the main sleep outcomes of interest.

## INTRODUCTION

Sleep state misperception (SSM) represents the discrepancy between measures of objectively recorded (i.e., with polysomnography (PSG) or actigraphy) and subjectively estimated (i.e., self-reported questionnaire) sleep. Such discrepancies can relate to the perception of one’s total sleep time (TST), wake after sleep onset (WASO) and/or the sleep onset latency (SOL)^1–3^. The presence of SSM is commonly found in individuals with chronic insomnia disorder^4^ and was the main feature of paradoxical insomnia in the second edition of the International Classification of Sleep Disorders (ICSD)^5^. Paradoxical insomnia was defined by the presence of subjective insomnia complaints (i.e., frequent difficulties falling asleep and/or maintaining sleep) not corroborated by objective sleep measures^2^. While the third edition of the ICSD^6^ revoked the previous classifications of different insomnia subtypes due to a lack of empirical evidence supporting these distinctions, SSM is still widely associated with the presence of insomnia^7^. Indeed, it has been shown that the severity of SSM is higher in insomnia disorder compared to other sleep disorders^3,8,9^, good sleepers^10–12^ and psychiatric disorders (e.g., mood disorders, trauma and stressor-related disorders)^2,13–15^. On average, individuals with insomnia disorder tend to overestimate their SOL and WASO and underestimate their TST, whereas good sleepers have been shown to either correctly estimate these sleep measures or overestimate their TST and underestimate their SOL and WASO^1,4^.

The assessment of SSM requires objective recordings of sleep and the subjective self-report of those same measures^1,4^. PSG is the gold standard in recording sleep objectively and allows a deeper investigation of the neurophysiological underpinnings of sleep (i.e., sleep architecture, brain oscillations)^16^. However, it is mostly performed in a laboratory setting and may be difficult to obtain during multiple consecutive nights. On the contrary, actigraphy, a wrist-wearable accelerometer device that records light and motion, can be used over multiple days at home for a more naturalistic investigation of an individual’s general sleep-wake patterns^17,18^. It is to be noted however, that although actigraphy has high sensitivity in detecting sleep epochs, it has low specificity when it comes to detecting wakefulness^19^. Actigraphy may also overestimate sleep time (longer TST, shorter SOL and WASO) in individuals with insomnia due to these individuals’ poor sleep hygiene (i.e., laying in bed motionless trying to fall asleep)^19,20^. Similarly, when te Lindert and colleagues (2020) assessed actigraphy scoring algorithms, they found that sleep and wake were less accurately detected in individuals with insomnia compared to good sleepers^21^. Furthermore, when studying individuals with various sleep disorders (i.e., periodic limb movement disorder, obstructive sleep apnea, upper airway resistance syndrome, insomnia disorder), it was found that objective SOL and WASO were of similar duration between PSG and actigraphy, but objective TST was more sensitive to the type of method used and may differ greatly between actigraphy and PSG^22,23^.

Multiple studies have used either actigraphy^14,21,24–29^ or PSG techniques^1,8–13,24,30–36^ in combination with self-reported sleep estimates to assess SSM and the effect of interventions in different populations. However, no studies have directly compared these two methods in the assessment of SSM. While most studies using PSG or actigraphy show that individuals with insomnia disorder tend to underestimate their sleep duration while overestimating sleep onset latency and wake duration, the degree of severity may vary based on the data collection method.

In sum, there is limited understanding on how the modalities of sleep recordings might impact the measurements of the severity of SSM and its characteristics, especially in insomnia disorder. Here, we aimed at providing a comprehensive assessment of SSM in individuals with and without insomnia disorder by comparing SSM (i.e., degree of misperception in TST, SOL, and WASO) extracted from one PSG night in-lab and multiple nights with actigraphy at-home, both complemented by subjective sleep reports. We hypothesize that individuals with insomnia disorder and good sleepers will perceive their sleep differently, specifically that those with insomnia will underestimate their TST and overestimate their SOL and WASO, while the good sleepers will perceive their sleep accurately (i.e., no mismatch between objective and subjective sleep) both in-lab and at-home. Due to the technological and environmental differences between in-lab (PSG/night review) and at-home (actigraphy/sleep diaries) methods, we expect that the severity of SSM will differ depending on the methods across groups.

## MATERIAL & METHODS

### Participants

Adults with an insomnia disorder and good sleepers were recruited via online and print advertisements posted locally in Montreal (QC, Canada), and from physician referral. Prospective participants were initially screened over the phone for inclusion and exclusion criteria. Next, a semi-structured in-person interview was conducted by trained research coordinators who administered the structured clinical interview for disorders (SCID-5) present in the 5th edition of the Diagnostic and Statistical Manual of Mental Disorders (DSM-5)^37^. This interview screened for the presence of insomnia disorder, major depressive disorder, general anxiety disorder as well as panic disorder, agoraphobia, alcohol use disorder, social anxiety disorder, obsessive-compulsive disorder, post-traumatic stress disorder, anorexia nervosa, and adjustment disorder. This interview also included demographic information and medical history. Potentially eligible participants subsequently underwent a screening polysomnographic (PSG) recording to rule out the presence of other sleep disorders (i.e., sleep apnea, periodic limb movement).

Participants meeting the DSM-5 diagnostic criteria for insomnia disorder for at least 3 months were included in the Insomnia (INS) group. Chronic insomnia is defined as self-reported dissatisfaction with sleep associated with difficulties initiating sleep (i.e., sleep onset latency greater than 30 min), difficulties maintaining sleep (i.e., wake after sleep onset greater than 30 min), and/or early morning awakenings (i.e., final awakening time earlier than desired by at least 30 min), for at least 3 times a week and for more than 3 months, combined with significant distress or impairments of daytime functioning^37^. Participants who reported being good sleepers and did not meet criteria for insomnia disorder, other sleep disorders or psychiatric disorders were included in the good sleepers (GS) group. Exclusion criteria were as follows: being aged younger than 18 years old, presence of a psychiatric condition other than depression and anxiety as listed above, medical conditions likely to affect sleep (e.g., epilepsy, multiple sclerosis, Parkinson’s disease, chronic pain, active cancer), other sleep disorders (e.g., moderate to severe sleep apnea defined by apnea-hypopnea index (AHI) > 15/h, restless legs syndrome, periodic limb movement disorder defined by an index during sleep > 15/h), a major cardiovascular condition or intervention, a recent severe infection, poor cognitive function (defined by a diagnosis of dementia or a Montreal Cognitive Assessment (MoCA) score <26^38^, shift work or changes in time zones over the past 2 months, and monthly use of recreational drugs or weekly use of prescription drugs that might affect sleep. All participants signed a written informed consent form before entering the study, which was approved by the Concordia University Human Research Ethics Committee. The present manuscript used the baseline data from participants who were included in projects published elsewhere^39^ and were registered as clinical trials (ISRCTN13983243 - doi. org/10.1186/ISRCTN13983243 and NCT04024787 - https://clinicaltrials.gov/ct2/show/NCT04024787). Participants included in the analyses had completed a PSG recording and the associated self-reported night-review, as well as at least 5 days of actigraphy recordings and their associated sleep diaries. A total of ninety-two participants were found eligible for the study, including 57 individuals with insomnia disorder (INS) and 29 good sleepers (GS). Their demographic information is presented in Table S1.

### Protocol

Within a month after the initial PSG screening, which also served as an adaptation night, all eligible participants underwent a complete sleep assessment. All participants wore an accelerometer device on the non-dominant wrist and filled a sleep diary every morning during two weeks at home. Then participants came back to the laboratory to complete a second PSG recording and, in the morning, filled a self-reported night-review about their PSG night as well as questionnaires about their usual sleep habits and self-reported sleep quality, including the Insomnia Severity Index (ISI) to assess the self-reported severity, nature, and impact of current insomnia symptoms^40^ (Figure 1).

**Figure 1.**
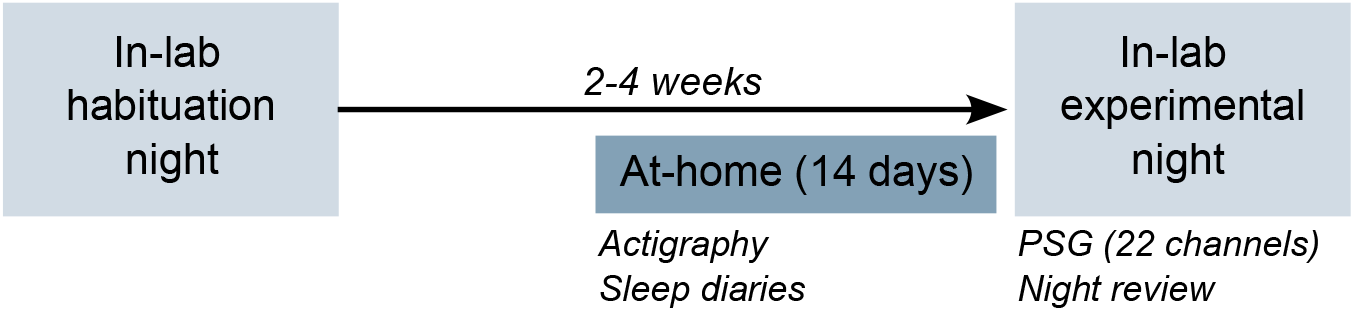
Study design.

### Measures

#### Collected at-home

##### Sleep diaries

Every day for 14 consecutives days, participants provided information about their sleep using the Consensus Sleep Diary^41^, either digitally or on paper. Their bedtime, sleep onset latency, number of nocturnal awakenings, time spent awake, sleep duration and sleep timing (bedtime and wake up time) were self-reported. Finally, participants rated their sleep satisfaction using a 5-point scale (1 being very bad, 5 being very good).

##### Actigraphy recordings

Participants were required to complete a sleep diary while wearing a wrist-wearable accelerometer device (Actiwatch 2 Philips Respironics) for 14 consecutives days. The device, worn on the non-dominant wrist, detects light exposure and measures motor activity in counts within a 30-s epoch (sampling rate: 32 Hz) which allow the assessment of the participants’ daily sleep/wake periods. Actigraphy data were analyzed automatically with the Actiware 6.0.9 analysis software and then, manually reviewed and edited in concordance with the sleep diary’s time in bed and time out of bed. When motion patterns did not match the times written in the self-report (over 30 min difference) we readjusted accordingly. Daily and averaged SOL, WASO, TST over the two weeks were extracted for each participant.

Over the two weeks, only recordings with a complementary sleep diary were analyzed (mean ± SD: all participants: 10.62 ± 2.52 days; INS: 10.60 ± 2.70 days; GS: 10.66 ± 2.16 days).

#### Collected in-lab

##### Night-review questionnaire

In the morning following each overnight PSG study, participants reported their subjective estimation of SOL, number of nocturnal awakenings, duration of WASO, and TST. Note that for measures of WASO in-lab, only 18 out of 29 GS and 41 out of 57 INS answered the subjective assessment of wake duration.

##### Polysomnographic (PSG) recording

Whole-night PSG recordings were used for all in-lab nights. PSG included EEG, EOG, EMG, and ECG. The 17 scalp electrodes (F7, F3, Fz, F4, F8, T3, C3, Cz, C4, T4, P3, Pz, P4, O1, O2, M1, M2) were placed according to the international 10-20 system. During the screening PSG (first night), periodic leg movements (legs EMG) and AHI (using thermistor, nasal cannula, thoracic-abdominal belts, and oximeter) were also computed to exclude other sleep disorders. The EEG signal was recorded with a Somnomedics amplifier (Somnomedics GmbH, Germany), sampled at 512 Hz. EEG recordings were referenced online to Pz, and for the offline analyses the EEG signals were re-referenced to the contralateral mastoids (M1, M2). All sleep scoring and analyses were conducted using the Wonambi python toolbox (https://wonambi-python.github.io). For each recording, two scorers blind to group allocation determined the different sleep stages (NREM 1, 2, 3, REM sleep and wake) according to the AASM rules for each recorded night of sleep^16^. From the scoring of the sleep architecture, we computed their SOL, WASO and TST.

##### Sleep state misperception measures

Measures of sleep misperception were extracted by comparing the degree of mismatch between objective measures (actigraphy and PSG) and subjective measures (sleep diary and night-review) collected at-home (actigraphy versus sleep diary) and in-lab (PSG versus night-review).

To assess the degree of misperception, we computed the Sleep Perception Index (SPI) which expresses the ratio between subjective and objective measures of sleep in a percentage value (subjective/objective*100). This provides a standardized measure of the perception of sleep duration (SPI-TST), sleep latency (SPI-SOL), and nocturnal awakenings (SPI-WASO)^12^. We extracted the in-lab SPI from the sleep measures from the PSG and the night review as well as the at-home SPI averaged over the daily SPI value extracted from sleep measures from the sleep diaries and actigraphy. Values around 100% indicate accurate perception while values <100% refer to an underestimation and values >100% refer to an overestimation of TST, SOL, and WASO. As a sensitivity analysis, we computed an alternative SSM measure by subtracting objective estimates from subjective assessments, resulting in SSM measures of deviations in minutes for TST, SOL, and WASO^3^ – see results in **Supplemental material**).

### Statistical analyses

Statistical analyses were performed using RStudio 1.2.50 (RStudio, Inc., Boston, MA) and R packages (e.g., car, emmeans, effsize, Rmisc, nparLD). The primary outcome variables were the SPI analyses of TST, SOL, and WASO. Secondary variables included the measures of objective and subjective TST, SOL, and WASO recorded in-lab and at-home. At-home measures reflected the average of multiples nights of recording for each participant while in-lab measures were extracted from a single night in-lab.

We used mixed-model analysis of variance (ANOVA) or the non-parametrical equivalent (Wald-Type Statistic) with Group (INS vs GS) as a between-factor and Method (in-lab vs at-home) as a within-factor. Post-hoc analyses were conducted using (paired and unpaired) *t*-tests or Wilcox tests. To assess the presence of misperception, paired t-test evaluated whether there was significant difference between subjective and objective TST, SOL and WASO. All analyses were conducted while adjusting for age and sex. When the measures were not normally distributed the statistical associations with age were assessed with a Pearson’s correlation and an unpaired Wilcox-test was used to assess sex effects. The normality of distribution was assessed with the Shapiro test and the homogeneity of variance was tested using the Levene test. Degrees of freedom were corrected according to the Greenhouse Geisser method when appropriate. The level of significance was set to a p-value of < .05 and p-values were adjusted for multiple comparisons (Benjamini-Hochberg/FDR correction). For significant results, both raw (p) and adjusted p-values (q) were reported when necessary.

## RESULTS

Eighty-six participants, including 57 INS and 29 GS completed a PSG recording and complementary self-reported night review, as well as at least 5 days of actigraphy recording and complementary sleep diaries. Overall, participants were middle-aged (44.56 ± 14.85 years old), and mostly female (74.42%). There was no difference in age (*t*-test *p*>.11) or biological sex (Fisher exact test *p*=.19) between groups. All participants completed the Insomnia Severity Index (ISI), which was higher in the INS group compared to the GS (*t*(85)=-18.55, *p*<.001; see **Table S1** for demographics).

### Influence of assessment method on objective and subjective sleep measures

First, we examined the potential effect that Method (in-lab and at-home: sleep diaries versus night review, and actigraphy versus PSG) may have on objective and subjective sleep variables (TST, SOL, WASO) separately. With regards to objectively recorded measures of sleep (PSG vs actigraphy), we found a Group by Method interaction (*F*(1, 85)=8.84, *p*=.003, *q*=.004) and main effect of Method (*F*(1, 85)=26.41, *p*<.001) for TST driven by shorter TST in-lab compared to at-home in the INS group only (*p*<.001). Meanwhile, we found a main effect of Method (*F*(1, 85)=13.53, *p*<.001) for SOL and a main effect of Group for SOL (F(1, 85)=5.17, *p*=.023, *q*=.046) and WASO (*F*(1, 57)=5.23, *p*=.022, *q*=.066) due to the INS group exhibiting shorter SOL (p=.004) but longer WASO (p=0.48) in-lab compared to at-home. In comparison, GS displayed no significant difference in sleep measures (SOL, WASO, TST) recorded by EEG (in-lab) and actigraphy (at-home; all *p*>.05) but revealed less WASO (*p*=.024) and longer TST (*p*=.003) in-lab than INS. SOL was significantly different between Group at-home only (*p*=.036 – **Table S2**). Meanwhile, in terms of subjective sleep measures (sleep diaries versus night review), there was a main effect of Group for TST (*F*(1, 85)=58.19, *p*<.001), SOL (*F(*1, 85)=37.28, *p*<.001), and WASO (*F(*1, 57)=21.91, *p*<.001) due to the INS group reporting shorter TST (all *p*<.001), longer SOL (all *p*<.001) and longer WASO (all *p*<.05) both in-lab and at-home compared to GS. We also found a main effect of Method (*F*(1, 85)=20.58, *p<*.001) for TST as both groups reported longer TST at-home than in-lab (GS: *p*=.033; INS: *p*<.001). However, there was no effect of Method on self-reports of SOL and WASO in both groups (all *p*>.05 – **Table S2**).

While we found no effect of age for objective sleep measures (all *p*>.05), we found that older participants in both groups tend to report shorter TST and longer WASO in-lab and at-home (all *p*<.05). We also found no biological sex difference except for female GS who reported a longer TST at-home compared to male GS (*p*=.045) but not in-lab.

### Assessment method and presence of sleep complaints impact sleep misperception

To investigate whether assessment of sleep misperception severity differed between at-home (actigraphy/sleep diaries) and in-lab (PSG/night review), we evaluated if the SPI of TST, SOL and WASO differed by Method and Group. To interpret this SPI, we used paired *t*-test between objective and subjective TST, SOL, and WASO where a significant difference indicates a mismatch between objectively recorded and subjectively reported measures.

For the SPI of TST, there was a main effect of Group (*F*(1, 85)=37.06, *p*<.001) which was driven by the INS group who exhibited smaller SPI than the GS group in-lab (*p*<.001) and at-home (*p*<.001). INS underestimated their sleep duration in-lab (objective TST vs subjective TST: paired t-test *p*<.001) and at-home (*p*<.001) while the GS group overestimated TST in-lab (*p*=.02) and at-home (*p*=.01). There was no main effect of Method (*F*(1, 85)=0.03, *p*=.86) nor an interaction between Group and Method (*F*(1, 85)=3.00, *p*=.08), suggesting no difference in the degree of misperception of sleep duration between at-home and in-lab across groups (**Figure 2**).

**Figure 2.**
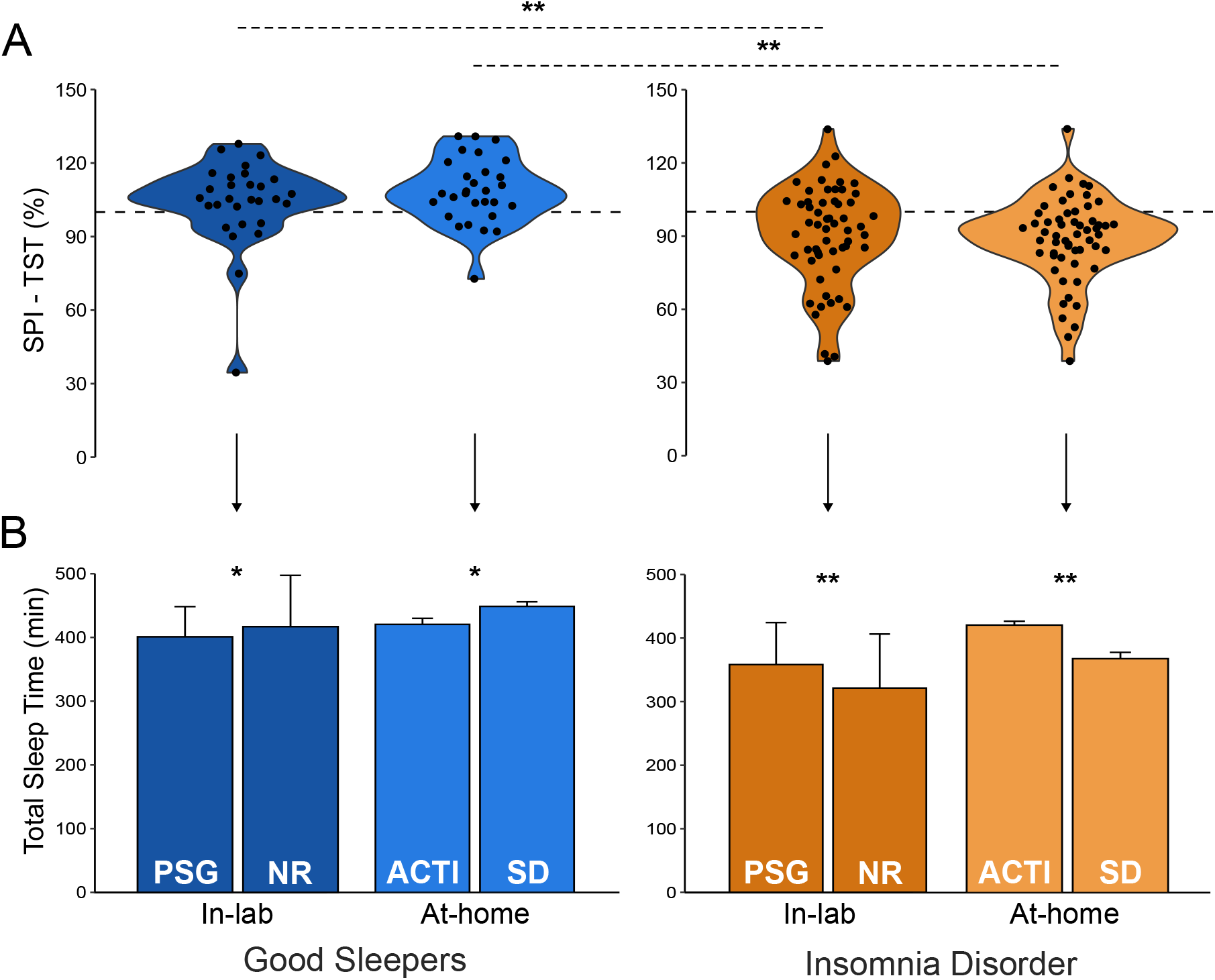
Sleep duration: sleep perception index and total sleep time across methods. (A) Mean and individual-specific sleep perception index of total sleep time (SPI-TST; %) as a function of Environment (in-lab: darker color; at-home: lighter color) for GS group (blue; N=33) and INS group (orange; N=59). Dot line at 100% represent accurate perception. (B) Mean (+SD) total sleep time (TST; in minutes) extracted from polysomnographic recording (PSG) and night review (NR) collected in-lab and mean (+SEM) TST extracted from actigraphy (ACTI) and sleep diaries (SD) collected at-home for GS and INS groups Asterisks represent significance (P): * <0.05; ** <0.001

For the SPI of SOL, there was no Group by Method interaction (*F*(1, 85)=0.64, *p*=.42) but there was a main effect of Group (*F*(1, 85)=6.68, *p*=.009, *q*=.01) and Method (*F*(1, 85)=16.78, *p*<.001). Indeed, while the GS group displayed correct perception both in-lab and at-home (all *p*>.05), INS exhibited higher SPI-SOL than GS at-home only (*p*=.01). The INS group overestimated their SOL in-lab (*p*<.001) and at-home (*p*=.007); however, the degree of misperception is larger at-home then in-lab (*p*=.001; **Figure 3**).

**Figure 3.**
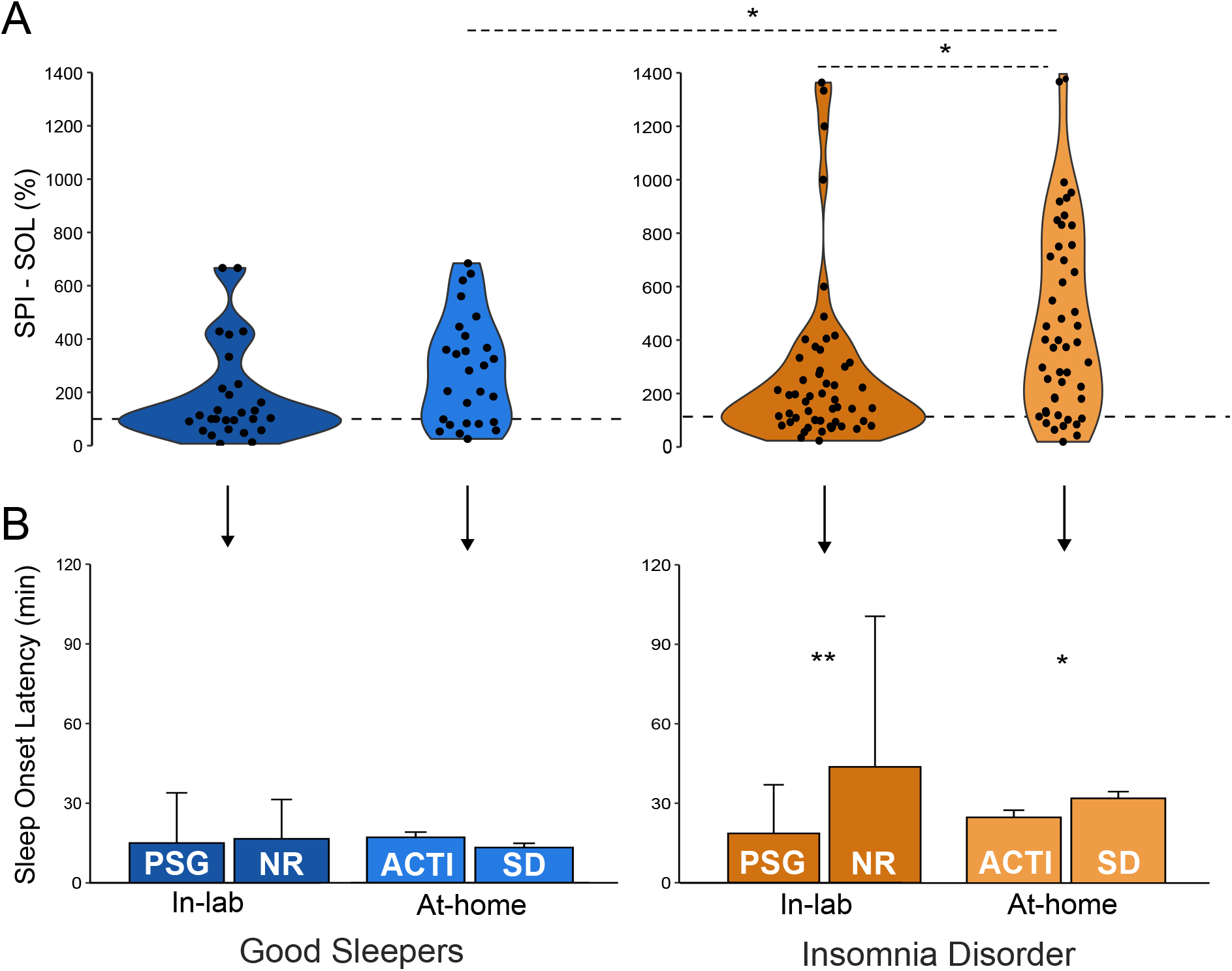
Sleep latency: sleep perception index and sleep onset latency across methods. (A) Mean and individual-specific sleep perception index of sleep onset latency (SPI-SOL; %) as a function of Environment (in-lab: darker color; at-home: lighter color) for GS group (blue; N=33) and INS group (orange; N=59). Dot line at 100% represent accurate perception. (B) Mean (+SD) sleep onset latency (SOL; in minutes) extracted from polysomnographic recording (PSG) and night review (NR) collected in-lab and mean (+SEM) SOL extracted from actigraphy (ACTI) and sleep diaries (SD) collected at-home for GS and INS groups Asterisks represent significance (P): * <0.05; ** <0.001

For the SPI of WASO, there was a Group by Method interaction (*F*(1, 56)=7.49, *p*=.006) with a main effect of Group (*F*(1, 57)=18.20, *p*<.001) and Method (*F*(1, 57)=8.57, *p*=.003, *q*=.005). Indeed, GS underestimated their WASO both in-lab (*p*=.009) and at-home (*p*<.001) while INS group exhibited higher SPI-WASO than GS at-home only (*p*<.001). As illustrated in **Figure 4**, INS overestimated their wake duration in-lab (*p*=.04) but correctly perceived it at-home (*p*=.47), suggesting that the SPI-WASO is dependent of the methods used in INS (*p*=.001).

**Figure 4.**
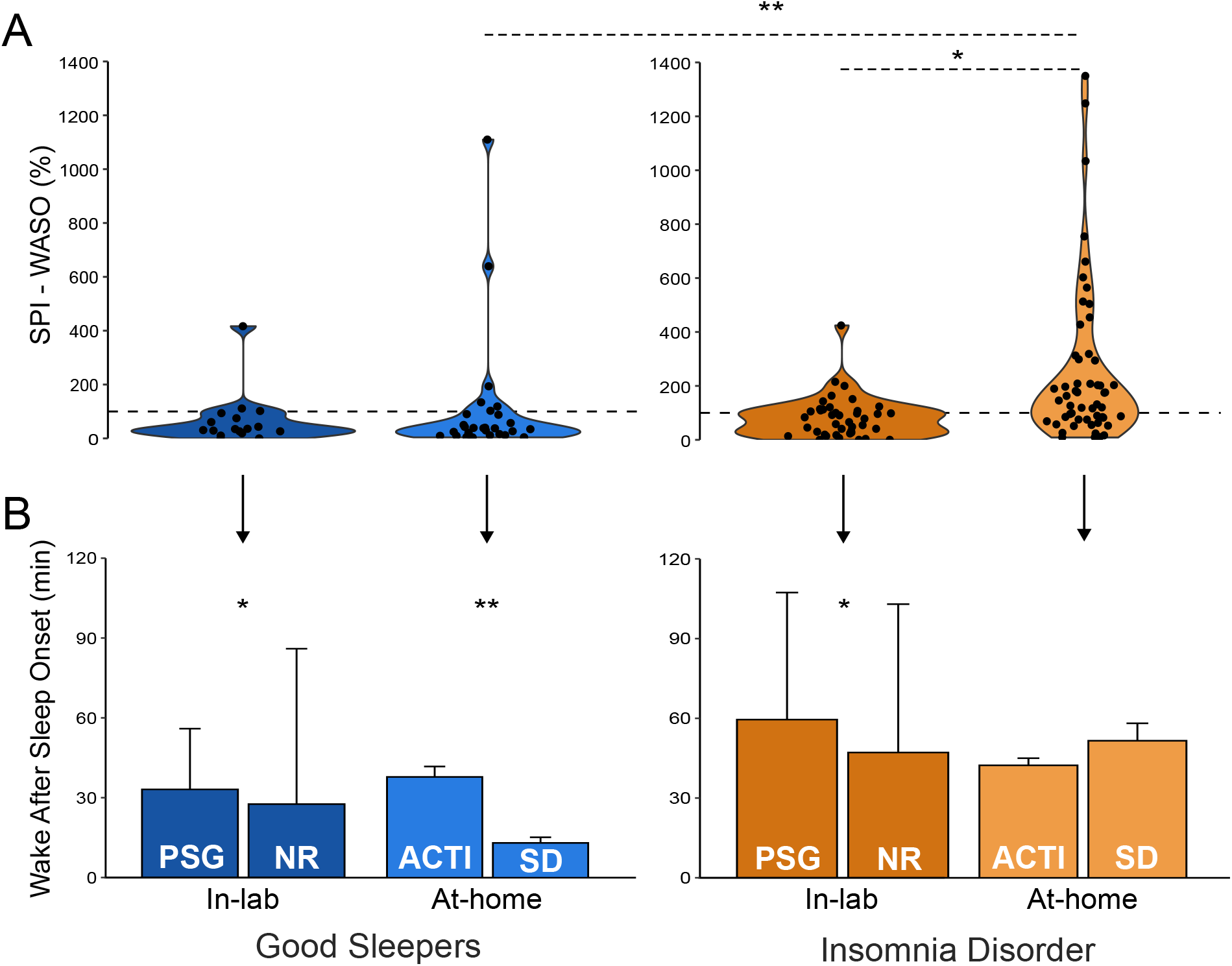
Wake duration: sleep perception index and wake after sleep onset across methods. (A) Mean and individual-specific sleep perception index of wake after sleep onset (SPI-WASO; %) as a function of Environment (in-lab: darker color; at-home: lighter color) for GS group (blue; N=20) and INS group (orange; N=43). Dot line at 100% represent accurate perception. (B) Mean (+SD) wake after sleep onset (WASO; in minutes) extracted from polysomnographic recording (PSG) and night review (NR) collected in-lab and mean (+SEM) WASO extracted from actigraphy (ACTI) and sleep diaries (SD) collected at-home for GS and INS groups Asterisks represent significance (P): * <0.05; ** <0.001

Interestingly, we found a significant correlation between SPI-TST at-home and SPI-TST in-lab (*r*=0.47, *p*<.001) in INS group. However, there was no association between SPI at-home and SPI in-lab for SOL and WASO across groups (all *p*>.05). While we found no effect of biological sex on the SSM variables (all *p*>.05), we found that across both groups, age was associated with increased degree of misperception. More specifically, older adults exhibited more severe overestimation of WASO in-lab (i.e., larger SPI-WASO; *r*=.28, *p*=.04) and severe underestimation of TST at-home (i.e., lower SPI-TST; *r*=-.38, *p*<.001) compared to younger participants. Association between age and TST at-home was mostly driven by the INS group (*r*=-.41, *p*=.002).

We also performed similar analyses using SSM measures in minutes, which is another computation of sleep misperception (report raw difference between subjective and objective measures in minutes) and found results mainly similar to SPI, especially for SOL and WASO (**Table S3**). Regarding SSM-TST, we found a difference between INS and GS groups, both in-lab (*p*<.001) and at-home (*p*<.001). However, we observed a Group by Environment interaction for SSM-TST (*F*(1, 85)=39.97, *p*<.001) driven by differences in the degree of misperception of sleep duration between at-home and in-lab across groups (**Table S3**).

## DISCUSSION

In this study, we provide a comprehensive methodological assessment of sleep state misperception (SSM - i.e., discrepancy between measures of sleep recorded objectively and subjectively perceived) in individuals suffering from insomnia disorder (INS) and good sleepers (GS). Particularly, we compared SSM measures extracted from one in-lab PSG night and multiple nights of actigraphy at-home, both complemented by subjective sleep reports. We found that GS and INS exhibit opposite patterns of sleep misperception. The degree of misperception in sleep onset latency (SOL) and wake duration (WASO) was larger in INS than GS, as has been shown previously^4,10,11^. However, we found that these measures were also dependent on the methods used to collect data, whether they were collected at-home or in-lab. Only sleep duration (TST) misperception was stable across methods within each group.

In the current version of the International Classification of Sleep Disorders^6^, the presence of insomnia subtypes (e.g., paradoxical insomnia, psychophysiological insomnia) is no longer recognized due to the inability to accurately distinguish between them^42^. In the example of paradoxical insomnia (defined by the presence of SSM), Castelnovo and colleagues (2019) found that more than 16 different cut-offs for SSM were used to characterize this subtype, which led to inconsistences in the prevalence of the diagnosis (14 to 64%)^1^. Currently, SSM is commonly accepted as a key process implicated in insomnia disorder rather than exclusively belonging to its own phenotype^7^. Indeed, in line with our results, most papers report that, on average, individuals with insomnia disorder overestimate their SOL and WASO and underestimate their TST, especially compared to good sleepers who have been depicted as accurate estimators of their sleep^4,10,11^. However, several studies using either actigraphy and sleep diaries at-home or PSG with night review in-lab also showed that there are interindividual differences in sleep misperception in both individuals with or without insomnia complaints and that the degree of misperception can vary quite widely in both groups^12,21^. Moreover, cognitive-behavioral therapy for insomnia (CBTi) - a multimodal psychological intervention aimed at modifying maladaptive thinking and behaviors that contribute to the perpetuation of insomnia^43,44^ – leads to significant improvements in the perception of sleep (i.e., reduce sleep misperception)^26,27,29,35,39^, suggesting that correcting sleep misperception may be important to the management of insomnia disorder.

Considering the increasing use of sleep misperception in both research and clinics, we aimed to provide a methodological evaluation of SSM at-home and in-lab. We found that only the measure of sleep duration misperception (SPI-TST) was robust across methods of data collection. In other words, whether it is collected on a single-night in-lab (i.e., PSG/night review) or over multiple nights at-home (i.e., actigraphy/sleep diaries), we observed a similar degree of misperception of sleep duration within both groups. Indeed, with both methods, we found that GS tended to overestimate their sleep duration while INS, on average, underestimate their sleep duration, supporting previous findings^4,10,11,31^. In the context of insomnia management, reduction in the misperception of sleep duration has been shown in studies using either recordings at-home (i.e., actigraphy/sleep diaries)^26,29^ or recordings in-lab (i.e., PSG/night review)^39^. Hence, these results suggest that the assessment of SPI-TST can be performed by either method depending on the scientific questions or the availability in methods in clinics and research laboratories. It is important to note however that, when SSM was calculated by subtracting subjective TST from objective TST (in minutes), the degree of misperception was dependent on the method used. Minute values for SSM was widely used in the past, specifically in the clinical definition of misperception in the diagnosis of paradoxical insomnia. However, Castelnovo and colleagues (2019) suggest that the use of a more standardized computation such as the ratio between subjective and objective measures of sleep may be more accurate in capturing SSM severity as the use of raw differences may bias inter-individual comparisons^1^.

Two of the main sleep complaints reported by individuals suffering from insomnia disorder are difficulties in falling asleep (i.e., long SOL) and maintaining sleep (i.e., long WASO)^6,37^. Interestingly, we found that SPI-SOL and SPI-WASO (as well as SSM-SOL and SSM-WASO in minutes) are dependent on the methods used, and importantly in the INS group only. Indeed, we observed that GS underestimated their WASO but correctly perceived their SOL in a similar manner when measured both in-lab and at-home (i.e., same degree of misperception across methods), suggesting that the assessment of SPI-SOL and SPI-WASO can be confidently interpreted using either method in good sleepers. In contrast, INS tended to overestimate their WASO in-lab but not at-home, and while they overestimated their SOL in both methods there was a greater degree of misperception at-home than in-lab. Such an inconsistency is not surprising as we found that INS objectively exhibited shorter SOL and longer WASO in-lab compared to at-home but did not report any difference in the subjective reports. The variability in the degree of misperception depending on the method of data collection might explain the disparity previously found in the diagnosis of paradoxical insomnia (14 to 64%)^1^. Indeed, depending on whether it has been collected at-home (actigraphy/sleep diaries) or in-lab (PSG/night review), data pertaining to SSM may have not reached the chosen cut-offs. However, beyond use of absolute cut-offs, when assessed as a continuum, a better estimation of SOL has been reported after CBTi in both studies using actigraphy/sleep diaries^27,29^ and PSG/night review^35,39^. In contrast, a change in the estimation of wake duration (WASO) following CBTi was reported by two studies using actigraphy/sleep diaries^26,27^ but it remained unchanged when assessed with PSG/night review in-lab^35^.

Our contrasting results concerning misperception in SOL and WASO emphasize the impact of the methods used to collect data (i.e., PSG vs actigraphy)^19,20,45^ as well as the environment (e.g., in-lab versus at-home)^46^ on the interpretation of the degree of misperception. From a methodological point of view, PSG provides precise measures of sleep and wake duration and thus degree of misperception when compared with subjective report. Moreover, multiple studies revealed that actigraphy has the tendency to overestimate sleep times and underestimate sleep latency and wake duration in insomnia disorder^19,20,45^, hence impacting the severity of misperception. However, it is important to consider the impact of environment. Indeed, it has been shown that multiple nights of actigraphy may be a more reliable measure of sleep characteristics compared to one night of PSG recording, especially in insomnia disorder which is characterized by higher night-to-night variability^21,47^. Finally, the combination of actigraphy and sleep diaries allows the study of sleep-wake rhythms at-home over several days without the confounding effect of sleeping in-lab (i.e., the first-night effect), which has been shown to alter sleep regulation with longer SOL and WASO^46^. However, this does not necessarily mean that actigraphy with sleep diaries are a better assessment of sleep misperception than PSG with a night review, as both methods bring complementary information about sleep. While the PSG and night review method can shed light on the neurophysiological signatures of sleep misperception^11,12,30^, actigraphy with sleep diaries allows a more naturalistic investigation of an individual’s sleep misperception patterns over multiple nights^21^. In the future, such limitations of the PSG related to the environment (i.e., in-lab) may be resolved by the emergence of using portable devices for monitoring sleep that can be used at-home over several nights^48^.

While we were able to compare information on sleep misperception extracted when using actigraphy/sleep diaries at-home versus PSG/night review in-lab in individuals with and without insomnia disorder, our work has some limitations. Although, we used a within-subject approach where all participants had data from the 4 methods used (i.e., single-night PSG and night review, multiples-night actigraphy and sleep diaries), our protocol did not allow comparison of sleep misperception measures during the same night of sleep, or the same number of nights in-lab compared to at-home. Indeed, the participants wore the actigraphy and filled the sleep diaries for two weeks and then spent the night at the laboratory. Thus, we can only infer the difference between the two methodological approaches of the same concept: SSM measured during one-night in-lab compared to the average of SSM measures over several nights at-home. It is important to note that we only assessed subjective WASO by asking the participants to retroactively recollect the previous night, which is not a precise and direct way of measuring subjective perception of wakefulness^34^. In addition, due to a non-compliance problem in answering questions on subjective WASO in-lab and at-home, the analyses of wake duration misperception were performed on a restricted number of participants in both groups (i.e., 18 GS, 41 INS). Finally, while we did not find biological sex difference in the severity of SSM, our sample was mostly composed of females (74.4%) which might hinder potential sex effect. Indeed, differences have been reported in objective and subjective TST, SOL and WASO between females and males,^49^ thus a more balanced sample size is warranted for future studies to properly characterize sex differences in SSM.

In summary, we found that only sleep duration misperception was reliably detected by both in-lab and at-home methods across good sleepers and insomnia disorder groups. Participants with insomnia disorder were more sensitive to the methodological approach employed as for the assessment of sleep misperception of sleep onset latency and wake time. Therefore, we recommend taking into consideration the methodology and environment depending on the population when assessing sleep misperception.

## Data Availability

All data produced in the present study are available upon reasonable request to the authors

## AUTHOR CONTRIBUTION STATEMENT

Conceptualization: AAP, AM, TDV; Funding acquisition: TDV, JPG; Project administration: FBP, LT, AAP; Data collection: AM, NW, KG, EMP, AAP, DS, LT, FBP; Data curation: AM, AAP, NW, EMP, KG; Formal analysis: AM, AAP; Methodology: AAP, AM ; Visualization: AM, AAP; Roles/Writing -original draft: AM, AAP,TDV; Writing - review & editing: AM, AAP, FBP, NEC, TDV, NW, EMP, KG, LT, DS, JPG

## ACKNOWLEDGMENTS

This research was supported by grants from the Canadian Institutes of Health Research (MOP 142191, PJT153115) to TDV and JPG. AAP has been funded by local awards (CRIUGM, PERFORM center, Concordia University) and the American Academy of Sleep Medicine. AM was funded by awards from the CRIUGM and Université de Montreal. EMP, NW and KG have been funded by awards from the Canadian Institutes of Health Research and local university awards (Concordia university, Université de Montreal). We acknowledge the contributions of the following lab members who assisted in participants ‘ recruitment, data collection and data preprocessing: Margie McCarthy, Emma Madigan, Ophelia Fontaine, Jennifer Suliteanu, Kazem Habibi, Brian Hodhod, Alex Hillcoat, Kajamathy Subramaniam, Rachel Hu, Loren Bies, Meaghan Pawlowski, Aminata Balde, Alexandros Hadjinicolaou, Victoria Yue, Laurence Vo Buu, Jonah Miller-Smith, Ilona Scellier-Dekens, Saba Sarani, Sampreet Arneja, Kimia Motevalli Bashi, Chelsey-Audrey Nkori and all the volunteers. We also thank our sleep technologists Madeline Dickson and Elinah Mozhentiy, and Liza Perez from the Clinique SomnoMed for their contribution to the setup of sleep recordings. Finally, we would like to thank the participants for giving their time and energy into this research study.

## ABBREVIATIONS

SSM: sleep state misperception PSG polysomnography
TST: total sleep time
WASO: wake after sleep onset SOL sleep onset latency
ICSD: internation classification of sleep disorders DSM diagnostic and statistical manual
INS: insomnia disorder GS good sleepers
AHI: apnea-hypopnea index SPI sleep perception index
CBTi: cognitive-behavioral therapy for insomnia

