## Supplemental material for "Methodological approach to sleep state misperception in insomnia disorder: comparison between multiple nights of actigraphy recordings and a single night of polysomnography recording"

### Supplemental results

Table S1. Demographics

| | All | | Insomnia (INS) | | Good sleepers (GS) | |
|---|---|---|---|---|---|---|
| Characteristics | Mean | (SD) | Mean | (SD) | Mean | (SD) |
| N | 86 | | 57 | | 29 | |
| Age | 44.56 | (14.85) | 46.33 | (13.73) | 41.07 | (14.85) |
| Sex | | | | | | |
| Female | 64 | | 45 | | 19 | |
| Male | 22 | | 12 | | 10 | |
| ISI | 12.02 | (7.81) | 16.84 | (4.32) | 2.55 | (2.78) |

*ISI, insomnia severity index*

Table S2. Posthoc differences in objective and subjective sleep measures according to Method and Group

| Objective measures | | | PSG | | Acti | | Method |
|---|---|---|---|---|---|---|---|
| | | | Mean | ± SD | Mean | ± SEM | p-value |
| TST | | GS | 400.95 | 47.38 | 419.85 | 9.42 | 0.31 |
| | | INS | 358.27 | 66.07 | 418.95 | 6.03 | <.001 |
| | Group | p-value | 0.003 | | 0.78 | | |
| SOL | | GS | 15.02 | 18.89 | 17.06 | 1.60 | 0.19 |
| | | INS | 18.62 | 18.35 | 24.58 | 2.51 | 0.004 |
| | Group | p-value | 0.18 | | 0.03 | | |
| WASO | | GS | 33.11 | 22.82 | 37.68 | 3.91 | 0.47 |
| | | INS | 59.52 | 47.80 | 42.42 | 2.70 | 0.048 |
| | Group | p-value | 0.02 | | 0.14 | | |

| Subjective measures | | | NR | | SD | | Method |
|---|---|---|---|---|---|---|---|
| | | | Mean | ± SD | Mean | ± SEM | p-value |
| TST | | GS | 416.90 | 80.42 | 447.98 | 7.25 | 0.03 |
| | | INS | 321.32 | 85.04 | 366.40 | 9.83 | <.001 |
| | Group | p-value | <.001 | | <.001 | | |
| SOL | | GS | 16.60 | 14.80 | 13.22 | 1.42 | 0.34 |
| | | INS | 43.72 | 56.84 | 31.70 | 2.37 | 0.41 |
| | Group | p-value | <.001 | | <.001 | | |
| WASO | | GS | 27.59 | 58.39 | 13.02 | 2.10 | 1.00 |
| | | INS | 47.13 | 55.83 | 51.70 | 6.58 | 0.91 |
| | Group | p-value | <.001 | | <.001 | | |

*PSG, Polysomnography; GS, Good sleepers; INS, Insomnia disorder; TST, total sleep time; SOL, sleep onset latency; WASO, wake after sleep onset*

Table S3. Group and Method effects on SSM computed in minutes

| Misperception (min) | | | Group | | Method | | Interaction | |
|---|---|---|---|---|---|---|---|---|
| | In-lab | At-home | F | p | F | p | F | p |
| **SSM-TST (min)** | | | 3.69 | 0.06 | 0.64 | 0.42 | 39.97 | <.001 |
| GS | 15.94 ± 63.46 | 28.13±10.48* | | | | | | |
| INS | -36.96±72.33†† | -52.55±9.47††** | | | | | | |
| **SSM-SOL (min)** | | | 0.85 | 0.36 | 7.55 | 0.006 | 0.64 | <.001 |
| GS | 1.58±14.64 | -3.84±2.60 | | | | | | |
| INS | 25.10±54.76 | 7.12±3.10†** | | | | | | |
| **SSM-WASO (min)** | | | 9.36 | 0.002 | 23.33 | <.001 | 11.82 | <.001 |
| GS | -6.38±54.55 | -24.66±4.01** | | | | | | |
| INS | -13.06±49.94 | 9.28±6.29†† | | | | | | |

**p<.05 within group differences; **p<.001 within group differences; †p<.05 between group differences; ††p<.001 between group differences*
